# Voices of the displaced: A qualitative study of potentially traumatising and protective experiences faced by refugee children

**DOI:** 10.1101/2022.07.26.22277918

**Authors:** Shaymaa Abdelhamid, Jutta Lindert, Joachim Fischer, Maria Steinisch

## Abstract

Refugee children experience potentially traumatic events that are distinct from the general population, yet current Adverse Childhood Experience (ACE) research addressing these specific adversities is scarce. This study uses qualitative interviews to identify potentially traumatising and protective experiences subjectively perceived as having an effect on the wellbeing of refugee children. Semi-structured interviews with 47 refugee parents and 11 children (aged 8-17) were conducted between November 2018 and January 2020 in the Rhine-Neckar region in Germany. Sampling was based on the official languages of the major nationalities seeking asylum in 2018, which were Arabic: Syria and Iraq, Farsi: Afghanistan and Iran, and Tigrinya: Eritrea. Informed consent from participants was obtained, and discussions focused on potentially traumatising and protective experiences before, during and after flight. Data collection was complete when data saturation occurred. Interviews were recorded, transcribed, coded and analysed using thematic analysis. We used the socio-ecological model to organise emerging themes that may arise at multiple levels and across all stages of migration. These experiences revolved around eight major themes including six themes reflecting on potentially traumatizing experiences: disruption, rejection, isolation, violence, impediments and affliction; and two themes that reflect on possible protective experiences: security/stability and connections. This study highlights important aspects to consider when examining refugee children’s experiences, such as addressing family dispersion, displacement, tough immigration and national policies as ACEs. It is increasingly important to identify these diverse experiences as the refugee population continues to grow and the increased prevalence of poor health outcomes in refugee children has been widely documented. Identifying ACEs specifically relevant for refugee children could contribute to understanding potential pathways and could further serve as a starting point for tailored interventions.

**Key messages:**

- Experiencing multiple adversities in childhood may lead to undesirable health and behavioural outcomes later in life. Current ACE literature does not capture refugee-related experiences, thus underestimating the full magnitude of the problems these vulnerable children could encounter.
- This qualitative study uncovered numerous potentially traumatizing and protective experiences specific to refugee children, such as displacement, cultural differences, insecure political climate as well as community support and access to education. It also suggests that a child’s wellbeing is influenced by multiple interacting components from the child’s family, community and society.
- Knowledge of new concepts significant to refugee children builds the foundation for developing an ACE questionnaire specifically for this vulnerable population – which could be a starting point for tailored interventions by identifying potential determinants for mental health and behavioural outcomes specifically relevant in refugee children.

## Introduction

By the end of 2021, 89.3 million people were forcibly displaced due to multiple emergencies, of which 36.5 million (41%) were children [1]. The humanitarian landscape is overwhelmed as new crises unfold. Within the last decade, the number of refugee children increased by 132% [2]. Refugees are fleeing war, violence, conflict or persecution to find safety [3]. Among refugees, psychological suffering may occur at each stage of migration caused by traumatic experiences in their home country and during flight, including the stress of adapting to a new life and culture in the host country. The latter, often unaddressed, can have a greater or equal effect than pre-flight stressors on the psychological well-being of refugee children [4, 5].

Research shows that undergoing potentially traumatic events in childhood – known as Adverse Childhood Experiences (ACEs) – is a potential pathway to social, emotional, and cognitive impairments leading to increased risk of unhealthy behaviours, violence, disease, disability, and premature mortality [6]. Previous ACE studies focus on adversities between the child and their family (abuse, neglect and household dysfunction) [7] and to some extent the community (bullying, discrimination and neighbourhood crime)[8, 9]. However, it is important to acknowledge that a range of factors on multiple levels influences a child’s health and development, as suggested by the socio-ecological model (SEM) [10]. In the SEM, the developing child is seen as being embedded in several milieus that affect their wellbeing, including family, community, and society [11]. Yet, not all studies consider such levels, often overlooking adversities associated with society (political climate, health, economic, educational, and social policies). Furthermore, adversities relevant to the refugee population such as war, displacement, or acculturation appear to be missing. These gaps highlight the need to acknowledge and explore the unique challenges refugee children face from all SEM levels.

To date, the majority of qualitative studies focus on adult refugee experiences. Of the studies that do discuss refugee children, many focus on a single migration phase [12], a single aspect of the SEM (e.g. refugee parenting behaviour) [13], or on internally displaced refugees (different experiences than resettling abroad) [14]. While important, these studies offer insights into a fraction of what refugee children experience, making it difficult to understand the full picture.

Equally important but similarly understudied is the identification of protective experiences that promote children’s development despite ACEs. The presence of healthy parents and nurturing environments, for example, are associated with fewer undesirable health outcomes [15]. In order to reduce further adversity, promote the children’s developmental abilities in areas such as resilience, discipline, stress-regulation, and empathy and encourage positive social, emotional and educational outcomes, refugee protective experiences must be identified.

In developing a clearer understanding of adversities refugee children encounter, and circumstances that could protect them, we must solicit the input of the individuals living these events. This study therefore explores the perceptions of refugee parents and children experiencing conflict, migration, and resettlement to uncover potentially negative and positive influences on the wellbeing of refugee children. In so doing, this study seeks to provide refugees with a voice, enabling a deeper understanding of sources of risk and resilience affecting refugee children’s health.

## Methods

### Setting and study population

The current study represents a component of a larger project entitled Beyond *Refugee Adverse Childhood Experiences* (BRACE) funded by the Deutsche Forschungsgemeinschaft (DFG, German Research Foundation) – GRK2350/1. BRACE is a mixed-methods project with two aims; the first involves qualitative interviews with refugees to inform item development for a questionnaire. The second seeks to establish the psychometric properties of the resulting questionnaire (reported separately). This project received ethical approval by the Medical Ethics Committee of the Medical Faculty Mannheim, Heidelberg University (approval no. 2018-610N-MA) and follows the COREQ guidelines for reporting results [16].

Interviews for the current study involved refugees in the Rhine-Neckar region in Germany. Recruitment was via convenience sampling through non-profit organisations, youth welfare facilities, and societies that organise civic engagement for refugee families. These five organisations granted access to locations where refugees lived/gathered. The target population included refugee parents and children (6-17 years) who were fleeing a conflict area and spoke either Arabic, Farsi, Tigrinya or German, the official languages of most asylum seekers in Germany when the study began [17]. No exclusion criteria were applied.

### Data Collection –Semi-structured Interviews

Participants were approached in person, introduced to the purpose of the study by the first author (SA, a female doctoral candidate), and invited to participate and ask questions; no relationship was established prior to study commencement. The interviews took place in either a room provided by an aforementioned organisation or in the participant’s home. SA, fluent in Arabic and English with intermediate German skills and fundamental knowledge of qualitative research, conducted the interviews. When needed, she was supported by a female Farsi-German interpreter, a female Farsi-English interpreter, a male Tigrinya-German interpreter or a female native German-speaking assistant. To offer support for participating children, a female child psychotherapist attended those interviews.

Participants were given the option to be interviewed either individually or in a group. The aim of this approach was to decrease refusals or withdrawals, since some participants were unable or unwilling to attend a group interview. To build trust with participants and ensure that they felt safe to talk openly, the study team decided not to collect any identifying data, thus preventing the opportunity for future contact [18].

Semi-structured interviews progressed between November 2018 and January 2020. An interview guide (appendix A) developed by SA welcomed participants, defined adversities and emphasised the importance of their participation. Child participants or parents speaking for their child were asked about potentially traumatising and positive experiences that may have occurred during each migration stage. Interview duration averaged 35 minutes (range: 15– 75 minutes). None ended prematurely and no incentives were provided. Data collection ended when theme saturation was reached.

### Data management and analysis

Written consent to audiotape interviews, take field notes and publish the results was given in each case, children under 16 assented and required a guardian’s consent. The recordings were transcribed by a professional transcription agency. Descriptors were removed to maintain anonymity. An independent native German-speaking collaborator translated German transcripts into English that were then checked for content accuracy by SA. Transcripts in English and Arabic were imported into MAXQDA 2018 (VERBI Software GmbH) for qualitative data management.

The transcripts underwent thematic analysis [19], SA developed and defined codes and assigned these to all transcripts in a descriptive manner. Using an online number generator, four transcripts were randomly selected for coding by a second independent individual to assess inter-coder agreement. Once coding assignment was complete, SA grouped the codes into possible emerging themes that were then reviewed and discussed within the research team until consensus was achieved. Following the SEM, codes were also organised to reflect influences on multiple levels. Codes with limited support (discussed by only a few participants) were documented for future exploration. Member checking to confirm accuracy of findings was not possible as no contact data was collected.

## Results

Thirty-six interviews with 58 participants were completed (Table 1). Eleven children (six unaccompanied and five accompanied) with a mean age of 14.6 years (range: 8-17 years) and 47 parents, mean age of 35.4 years (range: 23-63 years) participated. Most participants were female (45), the majority spoke Arabic and came from Syria (31), Iraq (6) and Palestine (4), followed by participants who spoke Farsi from Afghanistan (13), and Tigrinya from Eritrea (4). Many appeared to come from low socioeconomic backgrounds, evidenced by limited educational attainment (39) and unemployment (41). At the time of the interview, participants had spent an average of two years in Germany (range: 1 week to 4.5 years).

**Table 1.** Participant distribution in interview groups

|  | Number of participants per interview | Number of interviews | Interviews with adults | Interviews with children |
| --- | --- | --- | --- | --- |
|  | 4 | 3 | 3 |  |
|  | 3 | 2 | 2 |  |
|  | 2 | 9 | 8 | 1 |
|  | 1 | 22 | 13 | 9 |
| Total number of interviews |  | 36 | 26 | 10 |

Participants discussed numerous experiences, giving rise to eight emerging themes. We found evidence supporting six risk-related themes that appeared to represent *disruption, violence, impediments, affliction, isolation and rejection*; and two potentially protective themes: *security/stability* and *connections*. Experiences unique to refugee children were widely represented in the transcripts, not limited to the family milieu, but also appeared attributable to the community and society at all stages of migration. Representative quotations provided below for the respective themes and subthemes (*in italics*) reflect experiences mentioned by children themselves and by parents discussing what they considered to affect their child(ren).

### Theme 1: Disruption

One of the first themes to emerge from interviews revolved around disruption. A family-level disruption that several participants mentioned was *family bereavement (death of a loved one)*. Others perceived *family dispersion* as disruptive, since members of a family were scattered in different countries. Almost half identified that their families were separated either because a family member was at greater risk (e.g., forced military recruitment) or because the financial costs for traveling as a family were considered too high. This led some to send a child to a safe European country with the hope that family reunification applications for minors would be accepted faster.

Community life was also perceived to have been disrupted through displacement. Nearly all respondents recalled how they were forced to move several times, missing the opportunity to build bonds with others or establish roots:

> *“We have only been from camp to camp…we have been in camps for 3*
>
> *years. From Greece to…we went to Holland…they rejected us. The situation*
>
> *was very bad. I mean I have my [child], this little one,*
>
> *[s/he] is [an infant], [s/he]is psychologically unbalanced. I mean [s/he] doesn’t know the*
>
> *meaning of a home*.*”* - Syrian parent

Similarly, more than half of the children and parents commented on *disruption in education* before and during flight due to societal events related to an insecure political climate, forced school closures and problematic policies in transit countries.

### Theme 2: Violence

Almost all participants mentioned violence primarily at/within the community-level. Refugees expressed concerns for *extortion, exploitation, fraud, kidnapping, human trafficking, destruction, bombings, killing, fighting and physical harm*:

> *“The [foreign] guards caught us and beat us. They hit…you see my [child]?*
>
> *[S/he] was [an infant] when we left. The [foreign] guards hit [him/her], the*
>
> *situation is really… [shaky voice, crying]” - Syrian parent*

In addition, societal conditions encountered by many included surviving in the midst *of political insecurity, forced military recruitment, systematic violation of human rights, police/soldier brutality* (as mentioned by Speaker 4.1) and *militarisation*:

> *“The thing is there are the Taliban*…*We go through a lot of things…for*
>
> *example, I was at school for one day, not a week, because every day was*
>
> *war…I wanted to go…you could not go to school…there is no safe place when*
>
> *there is war*.*” - Afghan child*

### Theme 3: Impediments

Strong support existed for the perception of *impediments* to progress, especially in the form of *economic hardship*. Many participants reported losing their jobs and homes, making it difficult to afford necessities and escape safely. *Dangerous and long travel routes* were also given as examples of impediments to community life, with one unaccompanied minor taking four years to arrive in Germany.

*Immigration policies* were perceived as impediments as they complicated family reunification and often involved travel restrictions and long processing times for asylum applications:

> *“You [the government] are doing something good, for example, for the*
>
> *children, you are bringing [his/her parents for him/her], but what about*
>
> *[his/her] siblings? Are they not from the rest of [his/her] family? And they are*
>
> *minors. I mean the [child] has been waiting for [his/her parents] and family*
>
> *for three and a half years*.*”* Palestinian parent

*National policies* were recognised as another impediment with participants citing instances in which countries closed their border to prevent refugees from crossing, leading to their detention. Some described the Europe-wide fingerprinting scheme as a policy obliging refugees to return to the first European country in which their fingerprints were taken, impeding efforts in choosing their resettlement country.

### Theme 4: Affliction

Many respondents commented on *afflictions* in the form of *unfavourable psychological and physical health conditions* their children developed like asthma, skin infections and somatic symptoms from the stresses of migration. This is the only theme in which participants mentioned individual-level characteristics related to biological or personal history aspects. Many shared familial afflictions in the form of *parent’s distress*, recognising that their worry and fear was reflected in their children:

> *“The children only were afraid due to that stress that we had, [the*
>
> *parents]*.*”* - Afghan parent

Other forms of affliction included being homeless, living in a tent/container or living in overcrowded places under unhygienic conditions, or in other forms of *inadequate shelter*:

> *“We lived four years in a camp in Iraq* …*If it’s raining it would pour on us.*
>
> *When the weather is getting hot the tents burn, because of the electricity …*
>
> *the tents were burning*.*”* Iraqi child

### Theme 5: Isolation

Both parents and children mentioned matters regarding a sense of *isolation*. Several children missed in-person interactions and were *yearning for their relatives*. Strong support described how *cultural differences* due to multiple views, attitudes, languages, and traditions seem to elicit feelings of isolation for children trying to balance both cultures:

> *“There are huge differences between the way we raise our kids and our*
>
> *culture and between the way they raise their kids and their culture. Of course*
>
> *this will make us suffer. Our kids want to integrate “* Syrian parent

### Theme 6: Rejection

There was strong evidence in support of a theme centred on *rejection* in various forms including *discrimination* at the community-level and *refusal of asylum, revocation of refugee status or forced repatriation* at the societal-level:

> “They didn’t want to see so many Syrian people in Jordan. And that’s why
>
> we can’t do so many things. For example, this year, when I changed my
>
> school, we can’t talk to the Jordanian students. So they think we just have to
>
> have a Syrian school. We are separated. And you just think that, we are
>
> not normal.” - Syrian child

While the quotation above represents an example of discrimination refugee children faced during flight, many also described instances of discrimination pre-flight and a few mentioned post-flight prejudice.

### Theme 7: Security/Stability

Evidence for potential protective experiences was supported in comments about the theme of *security/stability*. For instance, *constructive parenting* where parents teach and model strength by encouraging patience, hope, gratitude, and depending on the child’s age either masking reality or explaining the circumstances was also viewed as important:

> *“I mean, when we lived in the tent and in the caravans, in the camps, a year*
>
> *and a half… I tell them it’s ok, this is a small phase and we will be patient.*
>
> *And we acclimatized and we got to know other people, refugees like us who*
>
> *were also unlucky and they were stuck in Greece, and we spent our days*…*”* -
>
> Syrian parent

Many children considered *educational value* as a form of *stability*, because they started believing that learning was important and it offered a pathway to a better future.

Strong evidence for this theme also emerged in reference to *community support*, which existed in different forms. Participants shared stories of community members providing them with practical (protection/transportation), informational (guidance) and emotional support as examples of *security*. Likewise, remarks about *stability* were made when participants discussed receiving material and practical (accommodation/translations) support that met their basic needs, for example, having a safe place to stay.

In addition, living in a *non-violent environment* with *basic human rights* and *social security* (family benefits, health insurance, social assistance, unemployment benefits and habitual residence) were societal-level conditions participants commonly mentioned. Such aids were perceived to inherently benefit and provide security for their children.

### Theme 8: Connections

Lastly, a common theme emerged around the value of connection with Germans, relatives nearby, people from their original culture, and forming true friendships:

> *“I had a ‘brother,’ to be honest and we were really fit on this path because he*
>
> *helped me and I helped him and so on*…*He was a good friend. And that was*
>
> *good, because it touched your heart so much. One does*
>
> *not think, one does not feel lonely in such situation…We were mutually healing for each other, so*
>
> *to speak*.*”* - Afghan child

### Subthemes with limited evidence

Other experiences were also discussed; however, evidence for each was limited as a few participants only mentioned them. Some recalled disruptive events such as *divorce, parent arrest* and a *missing parent* (whereabouts unknown). Similarly, violence in the form of *sexual abuse* and family *physical abuse* had weak evidence. A number of participants recalled impediments such as *lack of job opportunities and medical care* and a few noted afflictions such as changes in their *child’s development and behaviour, poor parental mental/physical health and parental drug use*. Additionally, evidence for circumstances that were interpreted as isolating such as *no family or community support and loss of network* had limited evidence. Lastly, rejection in the form of *neglect, bullying* and *rejecting own cultural customs* (e.g. arranged/child marriage) had limited support in the interviews.

Similarly, limited evidence also existed for perceived protective circumstances that could provide security/stability such as *presence of parents, financial stability, being rescued, fast resolution of asylum applications* and *opening of borders*. Also, only a few participants mentioned connections that maintained *ties to the child’s original culture and fitting in at school*.

## Discussion

Our analysis suggests that refugee children encounter many experiences at all stages of migration, on multiple socio-ecological levels. In this study, refugee ACEs appear to revolve around six themes whereas protective experiences revolve around two.

### Potentially traumatizing experiences

We can confirm that many ACEs reported in previous research among the general population (parent arrest, divorce, family death, parental neglect, physical abuse, sexual abuse and parental mental health [7]) are perceived as relevant for refugee children, too. Even though only a minority of participants mentioned some of these events, it seems worthwhile to examine the role of such adversities in the lives of refugee children in future research. In this study, however, participants appeared to emphasise stressors related to war, flight, and resettlement. Interviewees mentioned numerous experiences specific to the refugee population that, to our knowledge, have not been previously reported in ACE research.

Several experiences participants discussed are frequently mentioned in refugee literature, yet have not been accounted for when considering the types of adversities refugee children face. In considering each, we use the SEM to organise all emerging adversities, separated by the level at which they may arise and illustrate their potential interconnections (Figure 1).

**Figure 1.**
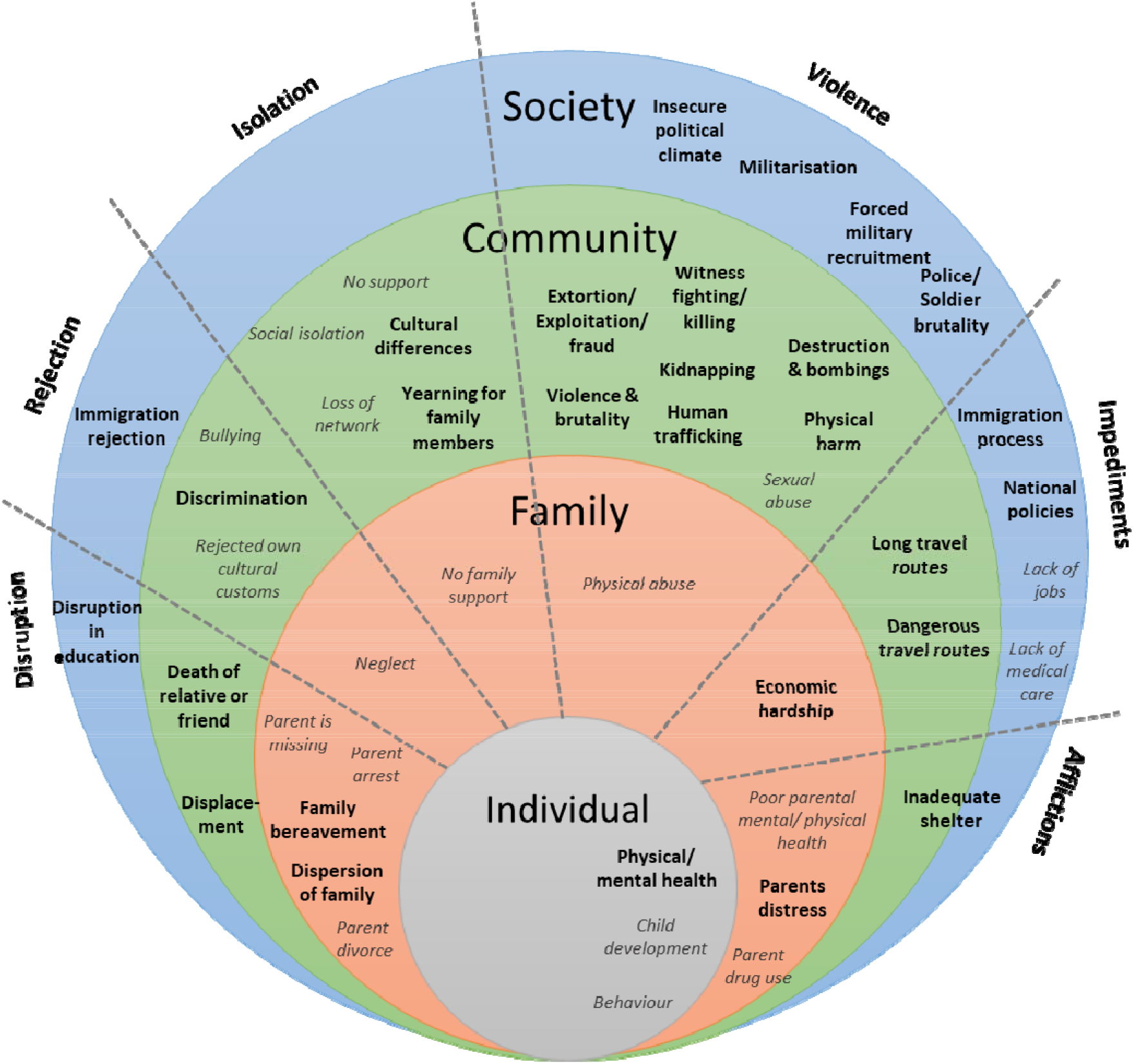
Six proposed themes and experiences* perceived as potentially traumatic (adapted from Dahlberg, L.L. et al. 2002 [20]) *Experiences in grey had weak evidence (mentioned by a limited number of participants)

Figure 1 represents the wide range of experiences throughout different levels of the SEM. We found evidence that interactions may occur and may have a direct or combined effect on the child’s wellbeing. For instance, refugees often experience *dispersion of family*, which may cause anxiety in children due to uncertainty regarding their parent’s whereabouts [21]. Additionally without parents’ physical presence, children tend to have behavioural problems, low academic achievement motivation and lack of self-esteem [22]. Similar consequences have been reported for *disruption of education*. Despite efforts to offer schooling to refugee children, accessibility depends more on the migration/asylum phase than on the child’s educational needs [23] leaving many children without education. A child’s critical thinking, confidence and stability are hindered by this societal-level disruption, consequently affecting their wellbeing [24].

Moreover, strong support existed for *displacement* in which community life is disrupted due to countless relocations. With resettlement efforts being slow (less than one percent of the 20.7 million refugees of concern to UNHCR in 2020 were resettled [25]) refugees are compelled to take long and dangerous routes with numerous obstacles sometimes leading to psychological and physical health consequences due to traumatic events during flight [21]. Displacement may prolong uncertainty, impede access to education and health care, hinder opportunities for parents to earn a sustainable living and impedes arrival in a safe/secure environment. This illustrates the perceived consequences of displacement which interacts with other community-level and family-level circumstances innately influencing a child’s wellbeing.

*Community violence* is sometimes experienced during displacement, and most often war is the reason behind it [3]. Interestingly, none of the children under 13 (ages eight, ten and twelve) discussed community violence, perhaps because they were too young when they fled their home country or they might not have personally experienced it. On the other hand, nearly all other participants regardless of their origin mentioned the many forms of community violence shown in Figure 1, suggesting that it is uniformly of high importance. Previous research has disclosed the relationship between this community-level adversity and individual afflictions, citing high levels of mental distress, depression, anxiety and post-traumatic stress disorder as unfavourable outcomes [26].

Additionally, strong support in our study existed for the perceived detrimental effects of *militarisation*, its negative impact seldom mentioned in other studies. Constant blockades preventing children from going to schools, preventing goods or people from entering or leaving, continuous interrogations and unwarranted raids of homes cause children to constantly feel in danger. This gap is critical because societal violence disrupts other societal constructs such as education and often triggers community violence in turn affecting children’s health and yielding mistrust in police and soldiers that are meant to keep citizens safe. Militant presence and police/soldier brutality are exposures rarely discussed in ACE or refugee qualitative literature.

*Economic hardship* is another element related to refugee children’s struggles. Many participants discussed how parental job loss and depreciation of the local currency caused difficulties in affording necessities and safe refuge. However, economic hardship goes beyond these implications. When parents become unemployed for a long time there are negative consequences for children’s academic achievement and some are encouraged into early marriage or child labour [27]. Our data suggest these difficulties are ones that refugees appear to face primarily before and during flight. The consequences of economic hardship have also shown an impact on the well-being of refugee parents, in turn potentially affecting the emotional health of their children [13], as previous research has established a correlation between parental health and children’s mental health [28].

The majority of child and adult participants also discussed *inadequate shelter* as a potential adversity. The UNHCR defines shelter as “a habitable covered living space that provides a secure and healthy living environment with privacy and dignity in order to benefit from protection from the elements, space to live and store belongings as well as privacy, comfort and emotional support” [29]. However, that was not what refugees described when discussing the different forms of shelter they lived in. Inadequate shelter is a potential health problem for refugee children both physically through the spread of diseases that may occur in overcrowded settings, and mentally due to stress/anxiety of living in an insecure environment [30].

Another adversity perceived as stressful on the community-level is *cultural differences* [31]. Participants stated this was mostly owing to the difficulties in communicating in German and juggling two cultures. Interestingly, refugee children were more likely to comment on cultural differences than refugee parents. This could be because children are more likely to be in contact with the host culture, due to school enrolment, and that the majority of the participants in this study were homemakers, thus limiting their contact with other people.

Previous work highlights the salience of *discrimination* that refugees perceive in their resettlement countries [12, 13, 32-34]. However, in this study, discrimination was more commonly encountered in participants’ home countries and en route, arising from historical conflicts such as the intolerance Kurdish people face in Iraq, Syria, and Turkey [35]. As with cultural differences, discrimination can affect children’s feeling of belonging, cause lower self-esteem, reduce their aspirations and negatively impact their mental and physical health [26].

Furthermore, discussions about *immigration rejection and policies* that impede refugee progress are known. The former causes children to feel rejected by society and are in constant fear and anxiety of another rejection or deportation [33]. The latter increases the duration of uncertainty, insecurity and distress [12, 34, 36]. Yet conversations about national policies such as the Dublin regulation [37] and negotiations such as the European Union-Turkey deal of 2016 [38] are often made without acknowledging the outcomes such policies might have on refugee children. Although the implementation of such national polices are meant to aid the humanitarian crisis, these policies are perceived by some refugees as forms of rejection because in certain instances they result in transfers of asylum seekers, detentions and travel restrictions.

In short, these interviews suggest patterns of interconnectedness among ACEs at all levels of the SEM, affecting the child’s wellbeing. We must therefore further examine these interrelated pathways that have been linked to undesirable outcomes.

### Potential protective experiences

Our data appears to reflect predominately the perceived hazards and risks incurred during the refugee experience that may be explained by negativity bias, a natural human tendency to attend more to negative than to positive experiences [39]. For that reason, the majority of possible positive experiences were only mentioned by a limited number of people, thus recognised as having weak evidence. Despite this, two positive themes emerged (Figure 2).

**Figure 2.**
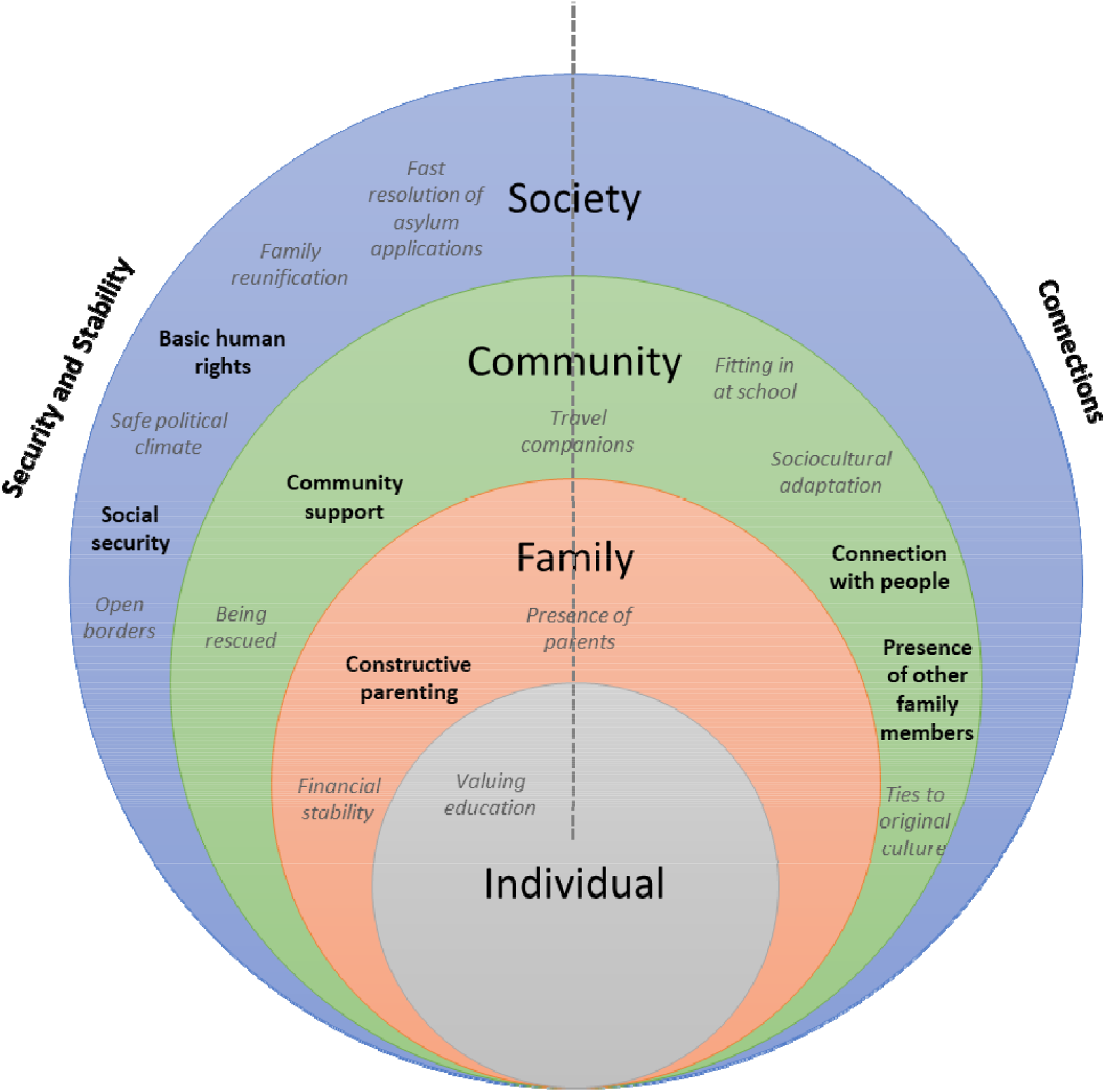
Two proposed themes and potentially protective experiences* (adapted from Dahlberg, L.L. et al. 2002 [20]) *Experiences in grey had weak evidence (mentioned by a limited number of participants)

It might seem that the experiences perceived as protective are merely the opposite of experiences perceived as potentially traumatic, which is true in some instances such as opening of borders and fast resolution of asylum applications. Open borders allow for safe passage of refugees instead of having to take dangerous routes or detention at borders, while the latter allows for refugees to settle into the new country, reduces stress regarding the acceptance of their application and allows them to be enrolled in schools all of which are good for the wellbeing of the child. However, potentially protective or adverse events can also be considered as extreme ends of a continuum [40]. For instance, constructive parenting is not simply the opposite of an absent parent; it is the presence of parents who are actively trying to provide their children with security and stability. Similarly, the majority of our interviewees vocalised the perception that they were now living in a place committed to ensuring their basic human rights including the right to life, freedom, work and education. This example of a safe and stable society is not merely the opposite of war or else it could have been found in transit/neighbouring countries. Refugees in Germany are provided with social security to be able to live with dignity as full, equal members of society [41], an important factor potentially contributing to stability and a sense of belonging in refugee children.

Similarly, community support in all its different forms builds important connections. One might argue that these connections are the opposite of isolation, yet the absence of isolation does not ensure a sense of connectedness. Support networks from the community help promote resilience by aiding refugees with their needs [42]. Likewise, connections either with relatives or other refugees allows for continuity of culture [43] while connections with people from the host community allows for sociocultural adaptation [32]. Additionally, school attendance offers children the opportunity to integrate with people from the community, possibly building friendships [44], while leisure activities such as football establishes relationships, enhances social integration and well-being [45].

### Strengths and limitations of the study

This qualitative study is the first, to our knowledge, to explore refugee children’s experiences at all stages of migration in different socio-ecological contexts and uncovers experiences not described in previous ACE work. Through the employment of interpreters, we were able to access refugees from a variety of ethnic backgrounds, making it possible to recognise emerging themes that were salient across cultures. Moreover, we aimed to ensure the rigor of this study by using qualitative methods such as using a semi-structured interview guide, audio recording, professional transcription, use of a computer software to organise codes, duplicate coding, and thematic extraction derived from the data via research team discussions. Another strength is that we interviewed refugee children about their own experiences, allowing them to add their own perceptions and voice matters that were important to them. Additionally, the combination of group and individual interviews, even though different data collection techniques, can be beneficial as they have the potential to increase knowledge of a phenomenon [46]. Group interviews offer opportunities to obtain a sense of the range of mutual views [47] while individual interviews provide more in-depth information [48].

Despite these strengths, a few limitations should be acknowledged. As common in qualitative research restraints on generalisability occur. The majority of interviews were with Arabic speaking participants. However, Arabic speaking refugees made up more than 36% of the refuge seeking population in Germany [17], and at the time of data collection the majority of refugees globally were from Syria [1]. The necessary use of interpreters might have resulted in comments that were under-translated or entirely lost in translation. However, Arabic is the first language of SA, who conducted the interviews, thus leading to fewer misinterpretations. Furthermore, the research team was aware of limitations caused by translations and tried to overcome this difficulty by employing interpreters that were bilingual and had interpreting experience with refugees. All interpreters were also coached prior to the interview regarding the importance of their work, the aims of this study and methods to avoid sections in the interviewee’s answer being untranslated/rephrased or interpreted. Similarly, an inability to confirm the presented findings through member checking raises the possibility of misinterpretation. However, member checking may also have a harmful impact on participants: recommendations exist that this process should either be avoided or implemented with caution when studying marginalised populations or traumatised participants [49] as re-engagement with the study topic and reading the presented findings might cause re-traumatisation [50]. Future research with refugees from other backgrounds, internally displaced people or refugees resettling in low-income countries may add to the findings presented in this study and shed more light on issues related to the generalisability of our findings.

## Conclusion

Refugee children clearly face multiple and ongoing challenges, yet numerous gaps in our understanding of the refugee child experience exist. Given continuous growth in the refugee population and previous research highlighting an increased prevalence of mental and physical health disorders among children associated with ACEs, it is increasingly important to understand the adversities affecting the wellbeing of refugee children and experiences that may be protective. This study adds new concepts to consider when examining ACEs in refugee children such as family dispersion, displacement, immigration and national policies. In addition, participants discussed constructive parenting, attaining basic human rights and having opportunities to build connections as potential protective experiences.

Refugee children’s encounters differ greatly from the general or even immigrant populations. Identifying and analysing their needs through qualitative research can add valuable insights to build a groundwork for future research and interventions but also for policy development. Drawing on this study’s results, several conclusions regarding potential interventions and policy re-evaluations can be drawn: For instance, re-evaluation of policies concerning children’s detention and reunion with their family is supported by our data, which is outlined by the Convention on the Rights of the Child (CRC) including the right for an education [51]. Other potential interventions supported by our findings include going beyond government based coverage of housing, medical care and minimum living expenses [41], for instance cash-based interventions [52]. Also, modifying emergency responses into more durable long-term solutions, like relocating refugees from camps to more private/suitable accommodation [53]; and parenting programs in refugee settings which have previously shown to have successful outcomes in reducing parental stress, improving parent-child interactions, thus improving development in young children [54].

In addition to the gaps in understanding the refugee child’s experience, screening and measurement tools to identify individuals that could benefit the most from targeted interventions are missing [55]. We anticipate our future work in the BRACE project will fill this gap.

## Data Availability

All data produced in the present study are available upon reasonable request to the authors.

## Acknowledgements

We gratefully thank all parents and children who shared their experiences with us, as well as the organisations for enabling access to the study population and the interpreters for their assistance in making this research possible. We also thank David Litaker MD, PhD (Center for Preventive Medicine and Digital Health, Medical Faculty Mannheim, Heidelberg University) for his valuable recommendations throughout the writing process.

