## Appendix A for "Voices of the displaced: A qualitative study of potentially traumatising and protective experiences faced by refugee children"

### Appendix A: Interview Guide

I want to start by thanking you for helping us with this research today. I welcome you to this meeting and want you to know that I am very happy to have each of you with us today. You are asked to help us better understand what you consider are potentially traumatic and positive life events that happen to refugee children. These negative experiences could be incidents that are incredibly upsetting, life-threatening or have an impact on your child’s physical/psychological well-being. While a positive experience is one that is pleasant and helpful to your child. Our objective is an important one. Identifying negative factors provides opportunities to intervene and treat these issues as well as prevent other negative outcomes from happening. Likewise identifying positive factors provides an opportunity to build on these positive factors and improve refugee children’s health and well-being.

Your input about these things is very important because you are an expert in your own life and know more about your friends and family than we do. Success will depend on your equal and full participation. Each of you here is an important group member, please feel free to share from your experience or experience of someone you know. There are no right or wrong answers, and I am not here to judge your comments in any way. I appreciate, the willingness of every one of you to fully share your ideas. The ideas which you generate in this meeting will become the basis for organizational planning for a questionnaire which will hopefully be used in the future to identify those in need and identify how to help them.

Do you have any questions?

Great, let’s get started.

Question 1: When you were in your home country…

What comes to your mind when you think about potentially traumatizing experiences for your children?

What were some events that happened that were upsetting or made you feel scared, or sad, angry or uncomfortable? These things can happen to any child not just you.

What are some positive experiences that you think protected your children?

What are some things that made you feel safe or happy that they were there?

Question 2: During your journey to Germany…

What comes to your mind when you think about potentially traumatizing experiences for your children?

What were some events that happened that were upsetting or made you feel scared, or sad, angry or uncomfortable? These things can happen to any child not just you.

What are some positive experiences that you think protected your children?

What are some things that made you feel safe or happy that they were there?

Question 3: As you resettle here in Germany…

What comes to your mind when you think about potentially traumatizing experiences for your children?

What were some events that happened that were upsetting or made you feel scared, or sad, angry or uncomfortable? These things can happen to any child not just you.

What are some positive experiences that you think protected your children?

What are some things that made you feel safe or happy that they were there?

These were the questions that I wanted to ask. Is there anything that you would like to add? Would you like to mention something that I did not ask you about?

Thank you very much for your participation, your contribution is greatly appreciated.
